## Supplementary material for "An Open One-Step RT-qPCR for SARS-CoV-2 detection": Suplementary information

### Supplementary Information

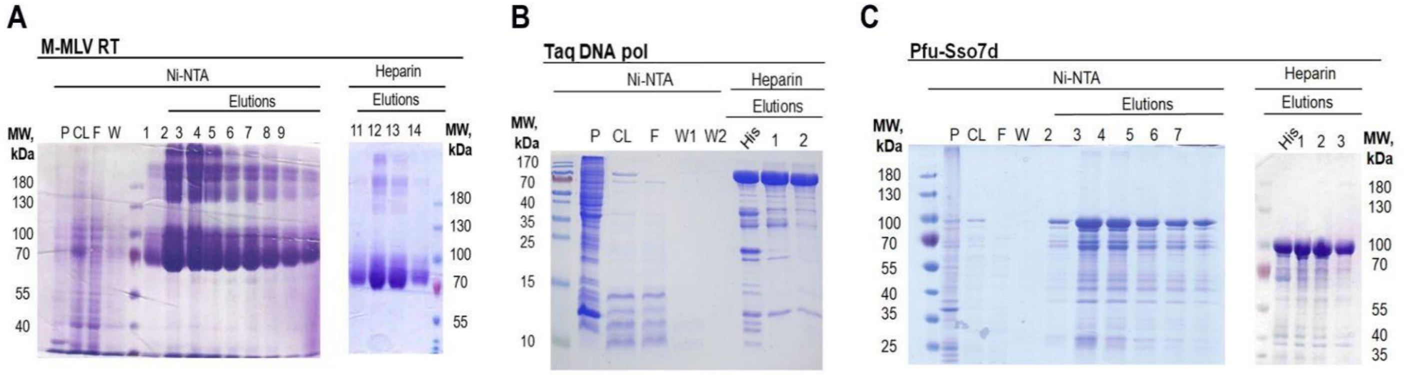


**Supplemental Figure 1. Purification of M-MLV RT, Taq DNA pol and Pfu-Sso7d enzymes.** SDS-PAGE of M-MLV RT **(A)** Taq DNA pol **(B)** and Pfu-Sso7d **(C)** purification. Samples were prepared by volume; 3 μL for pellet (P), clarified lysate (CL), and flowthrough (F); 12 μL for wash (W) and elution samples from the Ni-NTA and heparin purifications. The observed molecular weight of the purified proteins matches the one predicted based on their amino acid sequences: 80 kDa for M-MLV RT, 96 kDa for Taq DNA pol, and 100 kDa for Pfu-Sso7d. The His lane in the heparin purification panel in (B) and (C) corresponds to a pooled Ni-NTA purified protein sample before heparin purification. SDS-PAGE gels in (A) and (C) were made at 10% polyacrylamide (PA), whereas 12% PA was used for (B).

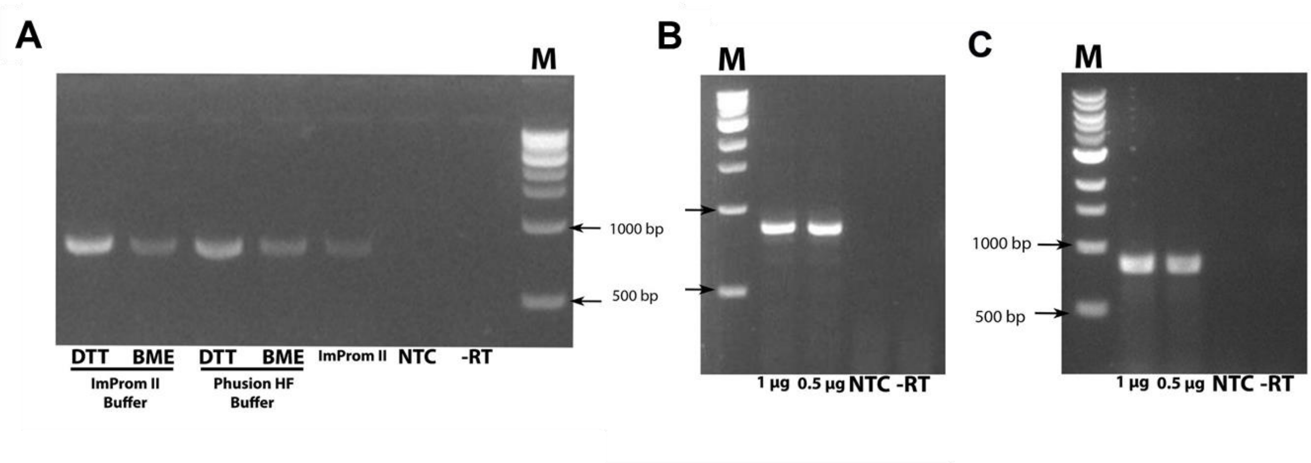


**Supplemental Figure 2. Homebrew M-MLV RT, Taq DNA pol and Pfu-Sso7d activity testing. (A)** Determination of M-MLV RT functionality in a conventional RT-PCR. A Two-Step RT-PCR was carried out using M-MLV RT for cDNA synthesis and Pfu-Sso7d for DNA amplification. ImProm-II 5X Reaction Buffer (Promega) and 5X Phusion HF Buffer (Thermo Scientific) were used. Additionally, dithiothreitol (DTT) and β-mercaptoethanol (BME) were tested as reducing agents. A positive control using ImProm II-RT in the 5X Reaction Buffer (Promega) was added. **(B)** One-Step RT-PCR using M-MLV RT for cDNA synthesis and Taq DNA pol for DNA amplification in a homemade 5X reaction buffer (composition described in Material & Methods) reaction buffer. Different amounts of total RNA treated with DNAse (1 and 0.5 µg) were used as a template. **(C)** One-Step RT-PCR using M-MLV RT for cDNA synthesis and Pfu-Sso7d for DNA amplification in the same conditions described in (B). The expected band size (~853 bp fragment), was obtained in all the samples. **NTC:** No template control. **-RT:** No RT control. **M:** DNA ladder 1 Kb (NEB).

**Supplemental Table 1. Dye-based RT-qPCR reaction mix using M-MLV RT and Pfu-Sso7d.**

| One-Step RT-qPCR (M-MLV/Pfu) | | |
|---|---|---|
| | **Volume (μL)** | **Final Concentration** |
| RNA sample | 5 | - |
| 10 μM Forward PCR primer | 0.6 | 300 nM |
| 10 μM Reverse PCR primer | 0.6 | 300 nM |
| 10 mM dNTPs | 0.8 | 400 nM |
| 5X Homemade Buffer | 4 | 1X |
| 100 mM DTT | 2 | 10 mM |
| 20X EvaGreen | 1 | 1X |
| Pfu Ss07d (0.6 mg/mL) | 0.5 | 15 ng/μL |
| M-MLV RT (0.02 mg/mL) | 0.125 | 0.13 ng/μL |
| Nuclease-Free Water | 5.375 | - |
| Total Reaction Volume | 20 | |

**Supplemental Table 2. Dye-based One-Step RT-qPCR cycling conditions.**

| Step | Temperature (°C) | Duration |
|---|---|---|
| 1 | 50 | 15 min |
| 2 | 98 | 2 min |
| **Repeat steps 3 and 4 by 40 cycles** | | |
| 3 | 98 | 10 s |
| 4* | 55 | 10 s |
| **Melt Curve** | | |
| 5 | 95 | 15 s |
| 6 | 60 | 1 min |
| 7 | 95 | 15 s |

*In this step fluorescence signal acquisition occurs (EvaGreen)

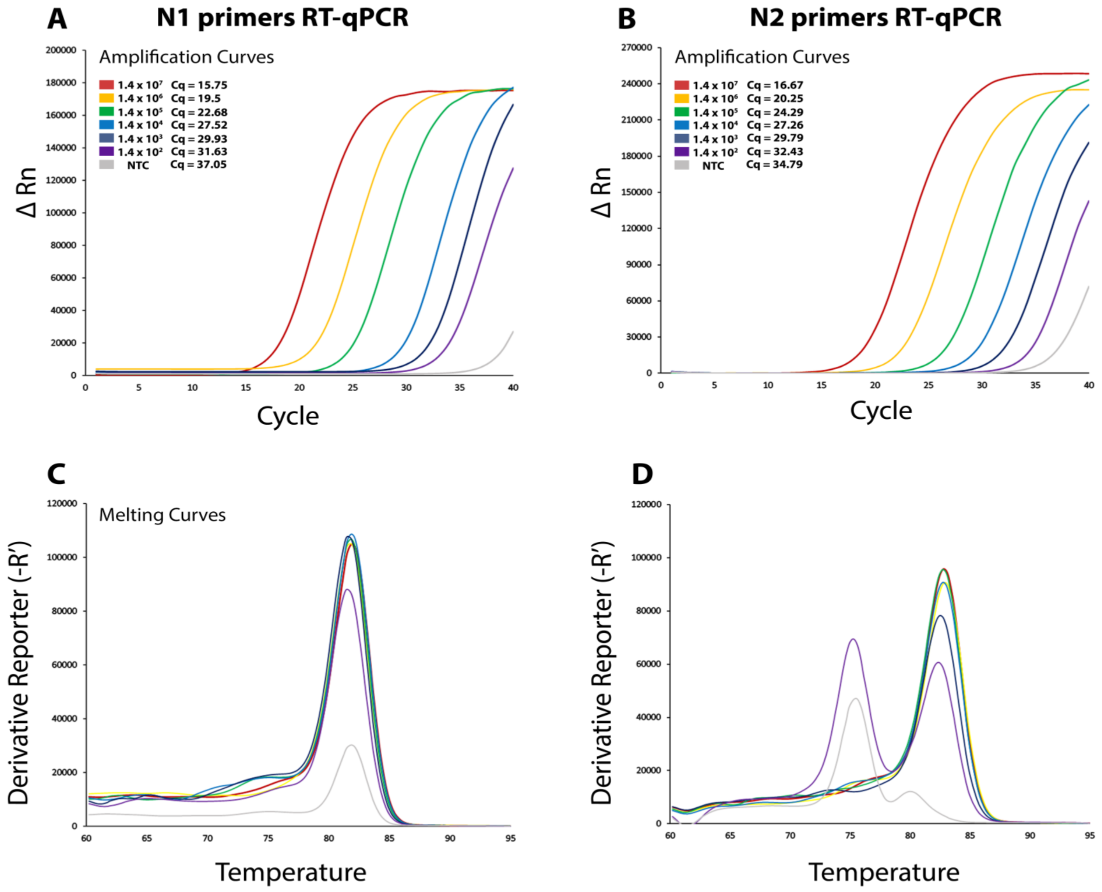


**Supplemental Figure 3: CDC SARS-CoV-2 N1 and N2 One-Step RT-qPCR assays performed with synthetic RNA using homebrew M-MLV RT and Pfu-Sso7d. (A-B)** Representative amplification curves using EvaGreen as DNA intercalating dye. Each curve represents a specific dilution of SARS-CoV-2 synthetic N RNA used as template: 1.4 x $10^7$ copies approximately (red line), 1.4 x $10^6$ (yellow line), 1.4 x $10^5$ (green line), 1.4 x $10^4$ (light blue line), 1.4 x $10^3$ (blue line), 1.4 x $10^2$ (purple line) and no template control (**NTC**, gray line). Characteristic Cq values are indicated on the upper left side of each panel in order to evaluate if amplification curves correspond to single amplicons (peaks); N1 and N2 melting curve analyses are shown in panels **C** and **D**, respectively.

**Supplemental Table 3. Comparative Cq data between a commercial RT-qPCR kit and an Open RT-qPCR method based on homebrew M-MLV RT and Pfu-Sso7d.**

| | | Commercial Kit | | | | Open RT-qPCR (M-MLV/Pfu) | | | |
|---|---|---|---|---|---|---|---|---|---|
| **# Sample** | **ID** | **N1** | **N2** | **RNAse P** | **Report** | **N1** | **N2** | **RNAse P** | **Report** |
| 1 | *4896* (+) | 13.78 | 13.60 | 27.13 | Positive | 13.16 | 13.98 | 26.79 | Positive |
| 2 | *5015* (+) | 14.26 | 14.66 | 26.60 | Positive | 13.16 | 13.73 | 27.10 | Positive |
| 3 | *5207* (+) | 16.46 | 16.33 | 30.67 | Positive | 14.9 | 17.56 | 29.76 | Positive |
| 4 | *5342* (+) | 16.53 | 16.45 | 28.95 | Positive | 16.06 | 17.21 | 31.58 | Positive |
| 5 | *5426* (+) | 16.68 | 16.28 | 30.62 | Positive | 16.22 | 20.3 | 29.25 | Positive |
| 6 | *4907* (+) | 17.31 | 17.95 | 28.18 | Positive | 16.59 | 17.83 | 27.58 | Positive |
| 7 | *5058* (+) | 18.98 | 18.82 | 26.88 | Positive | 19.33 | 35.79 | 27.02 | Positive |
| 8 | *5062* (+) | 23.89 | 24.00 | 25.90 | Positive | 22.88 | 23.79 | 25.42 | Positive |
| 9 | *5085* (+) | 23.91 | 23.89 | 27.48 | Positive | 22.29 | 22.96 | 27.62 | Positive |
| 10 | *5438* (+) | 25.01 | 25.61 | 31.28 | Positive | 24.4 | 25.52 | 30.70 | Positive |
| 11 | *4872* (+) | 26.61 | 26.81 | 31.19 | Positive | 25.04 | 27.07 | 30.88 | Positive |
| 12 | *4738* (+) | 28.24 | 28.30 | 27.54 | Positive | 28.2 | 29.43 | 29.88 | Positive |
| 13 | *5472* (+) | 30.15 | 30.86 | 29.62 | Positive | 28.91 | 29.99 | 29.09 | Positive |
| 14 | *4883* (+) | 30.44 | 30.87 | 28.25 | Positive | 30.35 | 31.97 | 28.89 | Positive |
| 15 | *5181* (+) | 30.86 | 30.29 | 29.54 | Positive | 28.99 | 29.29 | 30.66 | Positive |
| 16 | *4798* (+) | 31.98 | 31.19 | 27.92 | Positive | 31.43 | 34.43 | 27.50 | Positive |
| 17 | *5179* (+) | 34.07 | 34.60 | 31.58 | Positive | 33.29 | 34.23 | 30.51 | Positive |
| 18 | *4727* (+) | 34.18 | 33.60 | 31.14 | Positive | 32.2 | 31.96 | 31.95 | Positive |
| 19 | *4978* (+) | 34.92 | 35.16 | 28.83 | Positive | 32.3 | 32.62 | 27.29 | Positive |
| 20 | *5384* (+) | 35.58 | 34.26 | 26.10 | Positive | 33.21 | 32.77 | 26.86 | Positive |
| 21 | *2528* (+) | 35.84 | 35.04 | 26.10 | Positive | 34.45 | 35.36 | 26.40 | Positive |
| 22 | *5198* (+) | 35.99 | 35.21 | 31.66 | Positive | 31.75 | 31.59 | 30.72 | Positive |
| 23 | *5093* (+) | 36.04 | 35.55 | 26.03 | Positive | 33.15 | 33.99 | 26.11 | Positive |
| 24 | *5174* (+) | 36.11 | 35.62 | 25.57 | Positive | 33.82 | 32.42 | 26.44 | Positive |
| **25** | ***2514* (-)** | ND | ND | 26.36 | Negative | 35.03 | 37.91 | 26.42 | Positive |
| **26** | ***2529* (-)** | ND | ND | 27.40 | Negative | 35.2 | 34.66 | 25.32 | Positive |
| **27** | ***2486* (-)** | ND | ND | 28.96 | Negative | 36.62 | ND | 26.82 | Inconclusive |

| | | | | | | | | | |
|---|---|---|---|---|---|---|---|---|---|
| **28** | ***2513* (-)** | ND | ND | 27.26 | Negative | 34.47 | 35.15 | 26.63 | Positive |
| **29** | ***2489* (-)** | ND | ND | 25.33 | Negative | 36.38 | ND | 25.25 | Inconclusive |
| **30** | ***2487* (-)** | ND | ND | 27.44 | Negative | 37.17 | ND | 26.99 | Inconclusive |
| **31** | ***2477* (-)** | ND | ND | 28.85 | Negative | 35.34 | ND | 28.95 | Inconclusive |
| **32** | ***2473* (-)** | ND | ND | 25.90 | Negative | 37.32 | ND | 26.24 | Inconclusive |
| **33** | ***2517* (-)** | ND | ND | 28.32 | Negative | 37.32 | ND | 26.64 | Inconclusive |

Comparative Cq data for the TaqPath One-Step RT-qPCR kit and the dye-based open RT-qPCR reaction mix. Assigned sample number (# Sample) and clinical sample identifier (ID) are displayed. The clinical reports of the samples before they were re-tested by the two kits are also indicated in parentheses. The samples whose reports were altered are denoted in bold. **(-):** negative samples, **(+):** positive samples, **ND:** non-detected.

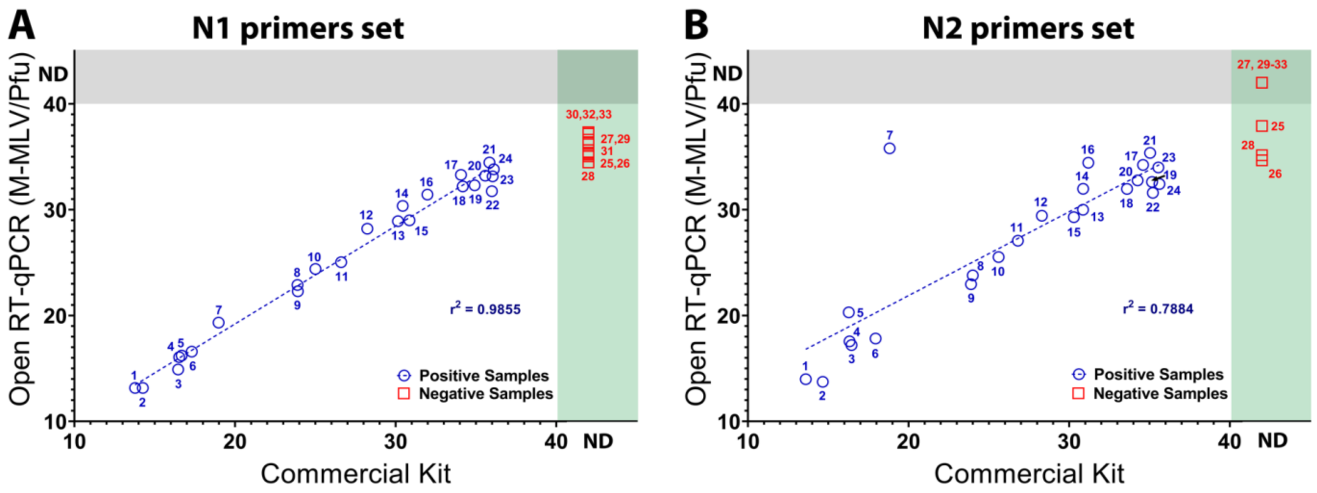


**Supplemental Figure 4: Negative samples cannot be successfully identified using the dye-based Open One-Step RT-qPCR reaction mix (M-MLV RT and Pfu-Sso7d).** Scatterplot of the Cq values of positive (blue circles) and negative (red squares) samples obtained by the commercial and open RT-qPCR reaction master mixes using the N1 and N2 primer sets (**A** and **B**, respectively). The numbers displayed in each sample match those displayed in Supplemental Table 3 (# Sample). If a Cq value was not detected in the sample, it appears in the ND area of the graph depending on whether this occurred in the commercial kit (green rectangle), the open dye-based kit (gray rectangle), or both (intersections between the rectangles). For each combination of primers, the linear trend of the positive samples is shown (blue dotted line) along with the corresponding value of $r^2$. **ND:** non-detected
